# Incidence and distribution of human leptospirosis in the Western Cape Province, South Africa, (2010-2019): A retrospective study

**DOI:** 10.1101/2022.01.05.22268774

**Authors:** Jacob Mugoya Gizamba, Lynthia Paul, Sipho Kenneth Dlamini, Jasantha Odayar

## Abstract

**Background:** Leptospirosis is an emerging zoonosis of global importance. In South Africa, the infection is an underreported public health concern, with limited information on its incidence and distribution. This study aimed to determine the incidence of human leptospirosis from 2010 to 2019 in Western Cape Province (WPC), and to compare the incidence based on seasonal and demographic factors.

**Methods:** A retrospective study was conducted with data on leptospirosis diagnoses by sex, age, season, and year in WCP obtained from the National Health Laboratory Services. With the provincial population sizes as the denominator, the incidence of leptospirosis was estimated and expressed as cases per 100,000 population. Negative binomial regression was used to estimate the effect of sex, season, and year on the incidence of leptospirosis.

**Results:** A total of 254 cases of leptospirosis were reported between 2010 and 2019, with the highest number of cases being in 2015. The annual incidence ranged between 0.15 and 0.66/100,000 population with a 10-year average incidence of 0.40/100,000 population. The incidence was higher among males than in females (0.55 vs. 0.25/100,000 population; incidence rate ratio (IRR) 2.2, 95% CI: 1.66,3.03). The 18-44 age cohort and had the highest average incidence (0.56/100,000 population), while the ≤17age cohort had the lowest incidence (0.07/100,000 population). The 18-44 (IRR 8.0, 95% CI: 4.65,15.15) and ≥45 (IRR 7.4, 95% CI: 4.17,14.17) age cohorts were more at risk of infection compared to ≤17age cohort. The average incidence of the infection was similar among seasons and there was no significant association between season and incidence of leptospirosis.

**Conclusions:** The results highlight that leptospirosis is an important zoonosis within the province disproportionately affecting males and the productive age demographic groups. These findings should enhance targeted prevention and provoke further investigation on the importance of environmental and socioeconomic factors on leptospirosis burden.

**Author Summary:** Leptospirosis is an emerging zoonotic bacterial disease of global importance. Despite its wide distribution, the disease is largely underestimated because its clinical manifestations mimic certain commonly known febrile illnesses such as malaria, influenza, hepatitis, and yellow fever. Leptospirosis burden in South Africa has been suggested to be moderately high however studies on the burden of the infection are lacking. This study sought to determine the incidence and trends of leptospirosis in the Western Cape Province, South Africa between 2010 and 2019. Overall, leptospirosis average incidence was 0.4 cases per 100,000 population (ranging from 0.15 to 0.66 cases per 100,000 population). Leptospirosis incidence was highest among male individuals and among those who were aged 18-years and above, however, the incidence did not differ by seasons. These estimates highlight that leptospirosis is an important zoonotic disease within the province and potentially disproportionately affecting males and productive age demographic groups. Therefore, this indicates the need for an all-encompassing One Health Approach to obtain all relevant information concerning leptospirosis distribution and risk factors in South Africa and in Africa at large to effectively enhance leptospirosis prevention strategies.

## 1. Introduction

Leptospirosis is an emerging, global zoonosis with an estimated 1.03 million cases and 58,900 deaths occurring annually [1–5]. The disease is caused by pathogenic spirochetes *Leptospira* species currently reported to comprise 66 different species with more than 300 serovars [6]. The infection in humans is associated with varying clinical manifestations ranging from a mild self-limiting febrile illness to severe illness characterized by dysfunction of multiple organs such as liver, kidneys, lungs, and the brain [7] potentially leading to pulmonary hemorrhagic syndrome and acute kidney injury due to Weil’s disease [5,8–10]. In addition, leptospirosis presents with symptoms that can mimic commonly known infections that cause febrile illnesses such as malaria, influenza, hepatitis, yellow fever, and viral hemorrhagic diseases among others [11,12]. Consequently, the infection has often been underdiagnosed and underreported [11]. Despite the life-threatening nature of leptospirosis, there is little published data on morbidity associated with the infection [4,5,13] contributing to its neglected status.

The risk of leptospirosis infection among humans occurs either through direct exposure to urine or aborted tissues of wild or domestic reservoir animals such as rodents and livestock/pets [4,14], or through indirect exposure such as contact with contaminated water, soil, and food [3,14]. In South Africa, a recent prison outbreak of leptospirosis was associated with exposure of inmates to rat urine in an overcrowded prison. Due to the predominant modes of transmission, leptospirosis is regarded as an occupational hazard affecting mainly farmers, sewer workers, veterinarians, and military personnel [11,15–18]. However, the transmission patterns are changing because there are increasing reports of leptospirosis in the general population. These cases are due to recreational exposures such as camping, kayaking, adventure travelling, hiking, cave exploration and other activities done in infected water [11,16,18,19]. Due to the diverse avenues through which humans can be exposed to the leptospires, occurrence of leptospirosis within the general population requires consideration [7,20].

Globally, tropical regions as well as resource limited settings are reported to have a higher burden of leptospirosis compared to temperate regions [5]. However, in Africa, few countries have data and reports on human leptospirosis [15,20] and research studies are scarce [2,5]. A previously published systematic review, of peer reviewed studies conducted in Africa, reported a prevalence of acute human leptospirosis ranging from 2.3% to 19.8% among hospital patients presenting with febrile illness and an estimated total of 750,000 cases per annum [15]. It was further concluded in this study that the morbidity of human leptospirosis in Africa is likely to be high relative to other global regions [15]. The need of more studies on the occurrence of leptospirosis in Africa was emphasized in another systematic review by De Vries et.al [20] to reliably understand the extent of the problem.

The incidence of human leptospirosis in South Africa has been suggested to be moderately high within the population [21–23]. Findings about the continued circulation of highly pathogenic *Leptospira* spp. among rodents suggests that the infection maybe an important underreported public health concern in the country [21,22]. A study conducted between 2009 and 2011 reported a seroprevalence that ranged from 9% to12.5% among all clinical samples sent to the NHLS for IgM ELISA testing from all over the country [23]. Environmental conditions in Western Cape South Africa, (the coastal, and temperate conditions with wet winters and warmer summers) have been suggested to favor transmission of pathogenic *leptospira* species *L. borgpetersenii* and *L. interrogans* traditionally associated with rats [4,21,22]. The most recent retrospective study conducted between 2005-2015, focused on patients presenting at a tertiary referral hospital in Western Cape Province (WCP), and a seroprevalence of 20% was reported [24].

Studies outside Africa have highlighted the influence of season, gender and age on the distribution and occurrence of human leptospirosis [25–27]. Such associations have scarcely been studied in Africa despite changes in climatic factors, demographic shifts, urbanization, and globalization [2,11,20,28–30]. The risk conferred by rodent infestation and overcrowding have been demonstrated in a recent leptospirosis outbreak that occurred in a prison in WCP, South Africa. [22]. Despite this information, studies describing leptospirosis occurrence in relation to demographic and seasonal factors are lacking in WCP.

To address these knowledge gaps, a retrospective study on human leptospirosis cases in the WCP, South Africa was conducted using data generated from diagnostic tests done in a public health care setting in Cape Town, South Africa. The study aimed at determining the incidence and trends of human leptospirosis over a 10-year period (2010-2019), with respect to year and seasonality and to describe the demographic characteristics of leptospirosis cases.

## 2. Methods

### 2.1 Study setting

WCP is located on the south-western coast of South Africa and is bordered by the Northern Cape and Eastern Cape Provinces. WCP has a population of approximately seven million inhabitants [31], three quarters of whom use public-sector healthcare services [32]. The province is approximately 129,462 km^2^ and has a population density of 45 inhabitants per Km^2^ [31]. Administratively, the province is divided into one metropolitan municipality (City of Cape Town) and five district municipalities (Central Karoo, Garden Route, Overberg, West Coast and Cape Winelands) [31]. The province has a diverse climate but is dominated by a Mediterranean climate with a cool, wet winter and a warm, dry summer. The inland daily maximum temperatures range from 20 ^°^C in winter to 32 ^°^C in summer and the mean annual rainfall is less than 380 mm.

### 2.2 Study design and study population

A retrospective study was conducted by using data provided by the data warehouse of the National Heath Laboratory Services (NHLS), on all human leptospirosis tests conducted at public healthcare facilities in the WCP, South Africa. The NHLS does serological testing on all serum samples of patients who are clinically suspected to be having a leptospirosis infection. Specimens from suspected patients are collected from different public healthcare facilities within the province and sent to a central NHLS laboratory in Cape Town. In this analysis, data included are from patients who had laboratory confirmed leptospirosis between 1^st^ January 2010 and 31^st^ December 2019.

### 2.3 Data collection

Human leptospirosis cases were considered for all ELISA IgM serological tests that were positive for Leptospirosis during the study period (2010 to 2019). ELISA IgM testing is the serological test conducted on all samples submitted to NHLS for leptospirosis screening and is sensitive in detecting new onset of illness [11]. Patient data extracted included the patient’s age, sex, year, date of test (year, month, and day) and the name of the health facility that submitted the specimen for testing.

The population sizes based on year, sex, and age in WCP for the 10-year period were extracted from the Western Cape Department of Health population circular H102 of 2020 [33]. This data was used as the denominator when calculating the incidence of leptospirosis infection.

### 2.4 Statistical analysis

All human leptospirosis cases were tabulated according to sex, age, season, and year of occurrence (2010-2019) and their frequency and proportions were calculated. The cases were not grouped based on their geographical location (district municipality) because the patient address information was not part of received data. Furthermore, the location of the health facility that submitted the specimen could not be assumed to be the same as the patient’s location because of the possibility of patients seeking healthcare or being hospitalized in a facility outside of their actual district of residence. The date when the test was done was used to assign the case to a particular year and season. Season was categorized as Summer (1^st^ December-28/29^th^ February), Autumn/Fall (1^st^ March-31^st^ May), Winter (1^st^ June-31^st^ August), and Spring (1^st^ September-30^th^ November). The variable year was conceptualized as a consecutive 12-month period from 1^st^ January to 31^st^ December, thus there were ten categories for year (2010 to 2019).

The incidence proportion of human leptospirosis for each year (2010-2019) was estimated and expressed as leptospirosis cases per 100,000 population. The incidence proportion by sex and age group as well as for each season of the year was also estimated. The Kruskal-Wallis Rank Sum Test was used to compare the average annual incidence between seasons of the year, age groups and between years. The Wilcoxon Rank Sum Exact Test was used to compare the average annual incidence between sex demographics. The negative binomial regression was used to estimate the effect of sex, gender, year of occurrence and season of the year on the incidence of human leptospirosis over the study period. This is a suitable regression model to use instead of the traditional Poisson regression in situations where modelling involves a count variable that is over-dispersed (the mean is less than the variance). The results were presented as incidence rate ratios (IRR) with 95% confidence intervals (CI). The cut-off value for statistical significance was 0.05. All statistical analysis was conducted using R software version 1.2.5033.

### 2.5 Ethical considerations

Ethical approval was obtained from the University of Cape Town Human Research Ethics Committee (UCT-HREC) (reference number: HREC REF: 303/2021). The Western Cape Provincial approval was granted by Groote Schuur Hospital Research Ethics Committee. The approval to use data from NHLS database was given by the NHLS Academic Affairs and Research office (reference number: PR2118953).

## 3. Results

### 3.1 Distribution of leptospirosis cases in WCP, 2010-2019

During the 10-year study period (2010-2019), a total of 254 cases of human leptospirosis were recorded by the National Health Laboratory Services (NHLS) (**Table 1**Error! Reference source not found.) for persons in the Western Cape Province, South Africa. Most of the cases reported had their specimen submitted from healthcare facilities located within the City of Cape Town Metropolitan (S2 Table 1). The highest number of cases was recorded in 2015 (42 cases, 16.5%) and lowest in 2012 (9 cases, 3.5%). When grouped by month of occurrence, the month of March (30 cases) had the highest number of cases over the 10-year period while September (16 cases) had the lowest number of the cases (**Fig 1**). When grouped into seasons, more cases were observed in fall season (75 cases), followed by summer, spring, and winter (64, 62, 53 cases respectively) (**Table 1**). More males were infected than females, with the males accounting for 173 (68.1%) cases compared to 81 (31.9%) cases who were females, and the overall male to female ratio was 2.136:1 (**Table 1**). The median age of the observed cases was 37.0 (Inter Quantile Range (IQR) 28.0-48.0) years, with a mean age of 38.12 years. Overall, 163 cases (64.2 %) were in the 18–44-year-old age group, 78 cases (30.7%) were in ≥ 45-year-old age group while 13 cases (5.1%) were in the ≤17-year-old age group over the 10-year study period (**Table 1**).

**Table 1:** Distribution of leptospirosis cases by age, sex, season, and year of occurrence in Western Cape Province South Africa (2010-2019)

| **Variables** | **All Cases (n=254)**<br>**n (%)** |
| --- | --- |
| **Age (years)** median (IQR) | 37.0 (28.0,48.0) |
| **Age group** |  |
| ≤17 | 13 (5.1) |
| 18-44 | 163 (64.2) |
| ≥ 45 | 78 (30.7) |
| **Sex** |  |
| Male | 173 (68.1) |
| Female | 81 (31.9) |
| **Season of the Year** |  |
| Summer | 64 (25.2) |
| Fall | 75 (29.5) |
| Winter | 53 (20.9) |
| Spring | 62 (24.4) |
| **Year of occurrence** |  |
| 2010 | 34 (13.4) |
| 2011 | 19 (7.5) |
| 2012 | 9 (3.5) |
| 2013 | 12 (4.7) |
| 2014 | 28 (11.0) |
| 2015 | 42 (16.5) |
| 2016 | 37 (14.6) |
| 2017 | 22 (8.7) |
| 2018 | 31 (12.2) |
| 2019 | 20 (7.9) |

IQR: Inter Quantile Range; n: frequency.

**Fig 1:**
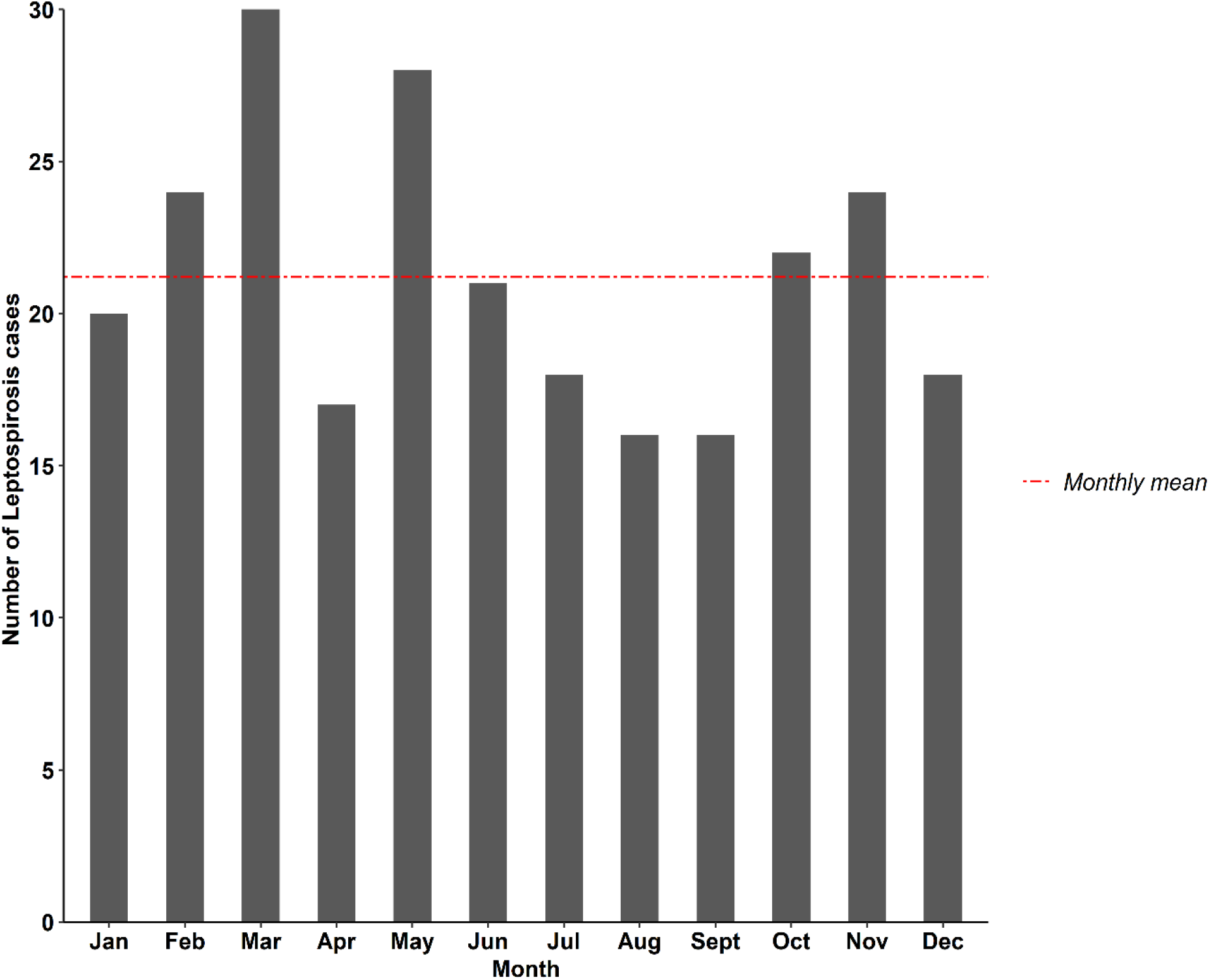
**Monthly distribution of leptospirosis cases in Western Cape Province over a 10-year period (2010-2019)**

### 3.2 Incidence of leptospirosis in the WCP, 2010-2019

The incidence of leptospirosis fluctuated widely across the 10 years. The annual incidence ranged from 0.15 to 0.66 cases per 100,000 population, with an average annual incidence of 0.40 cases per 100,000 population. The annual number of cases and the incidence are plotted against the year of occurrence to show the trends across the years (**Fig 2**). The incidence followed a downward trend between 2010 (0.59 cases per 100,000 population) and 2012 (0.15 cases per 100,000 population). However, there was an increase in the incidence from 2013 (0.20 cases per 100,000 population) which peaked in 2015 (0.66 cases per 100,000 population). This was followed by a gradual decrease in the incidence between 2015 and 2019 (0.66-0.27 cases per 100,000 population) except for year 2018 where the annual incidence was higher than the one observed in 2017.

**Fig 2:**
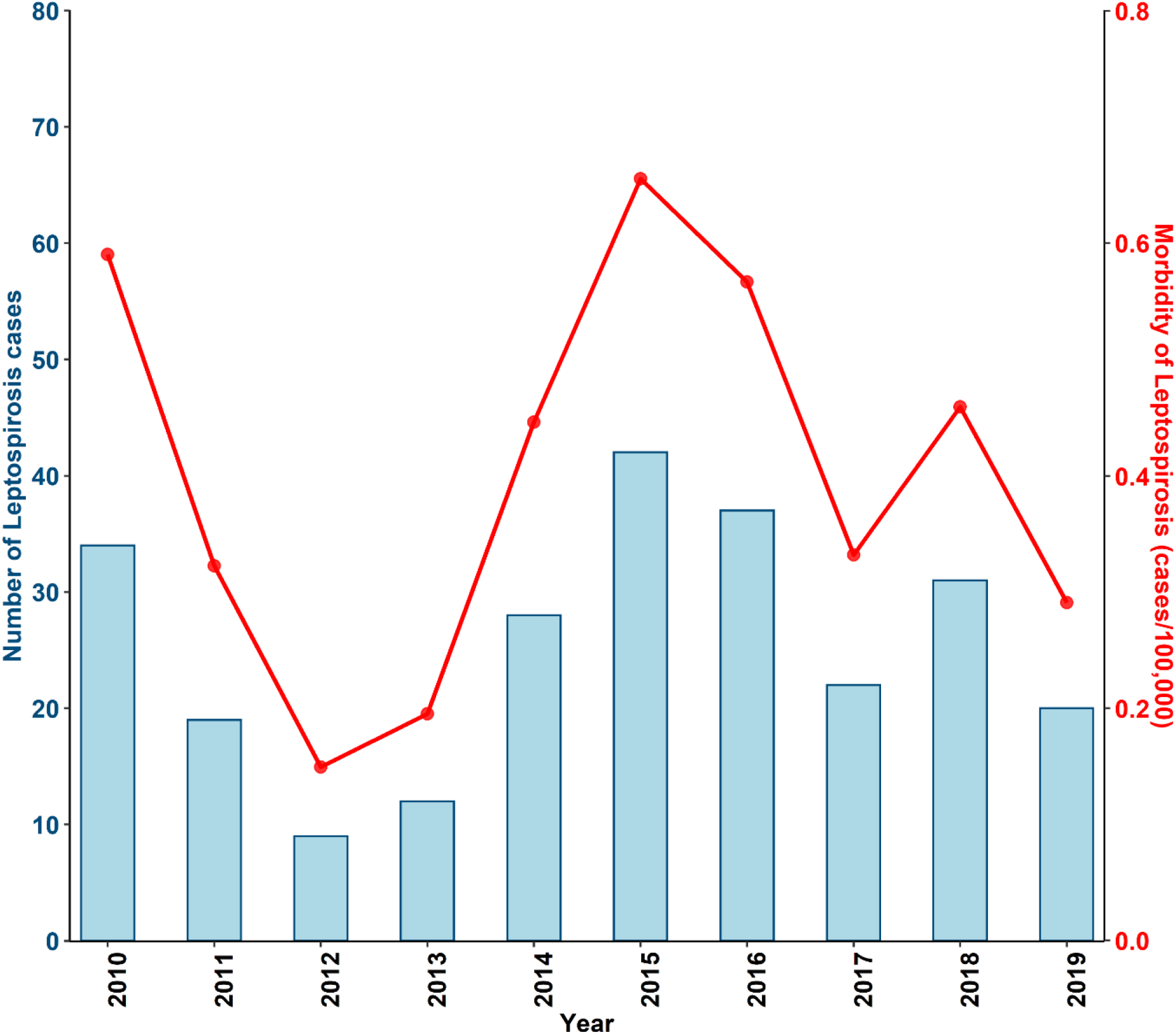
**The number of cases and incidence of leptospirosis per year (2010 to 2019) in Western Cape Province, South Africa.**

Throughout the study period, the incidence was highest among males compared to females (**Fig 3**), with the average incidence among males being 0.55 cases per 100,000 population and 0.25 cases per 100,000 population among females (p-values=0.004). During the entire study period, the incidence was highest in the 18-44-year-old and ≥ 45 years-old age groups compared to the ≤17-year-old age group (**Fig 4** and **Error! Reference source not found**.). The average incidence differed substantially between the age groups with the highest average incidence being among the 18-44-year-old age group (0.56 cases per 100,000 population) over the 10-year period, followed by ≥ 45 years-old age group (0.49 cases per 100,000 population) and the ≤17-year-old age group had the lowest incidence (0.07 cases per 100,000 population) (**Error! Reference source not found**.).

**Fig 3:**
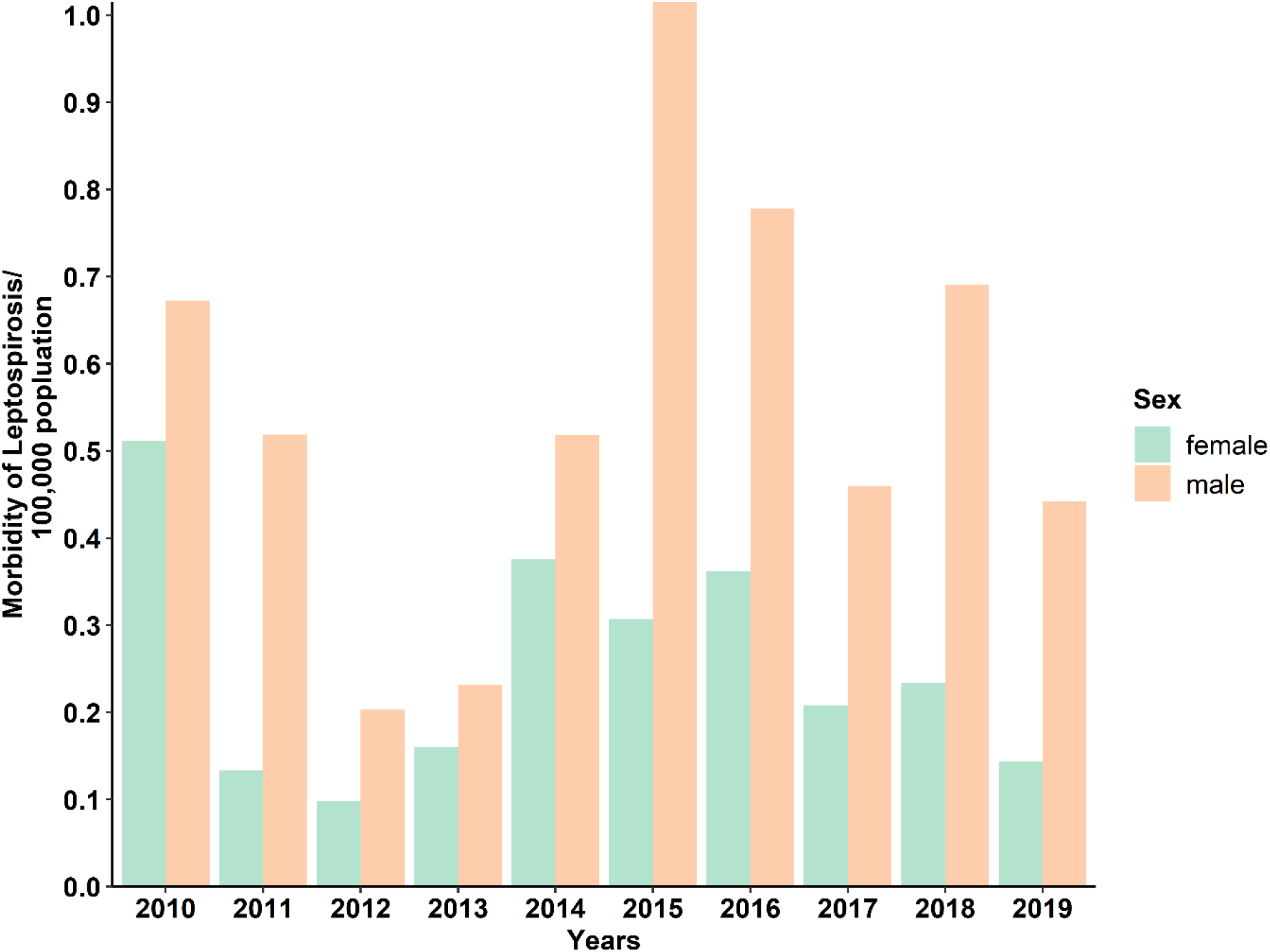
**Incidence of leptospirosis in males and females from 2010 to 2019 in Western Cape Province.**

**Fig 4:**
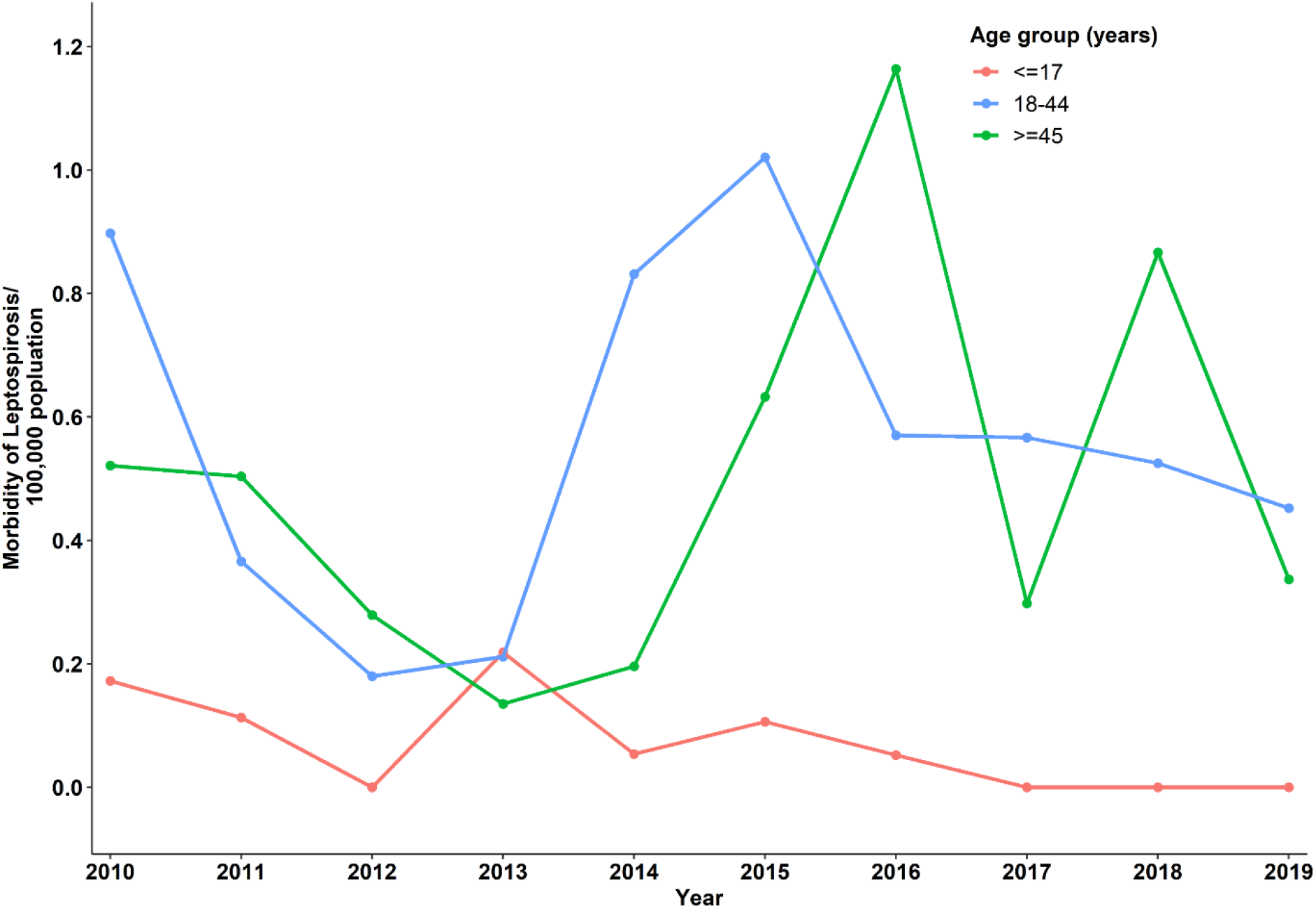
**Incidence of leptospirosis by age group from 2010 to 2019 in Western Cape Province, South Africa.**

The results indicate that on average, the annual incidence was higher during the fall season (0.12 cases per 100,000 population) and lowest in the winter season (0.08 cases per 100,000 population) over the 10-year period (**Table 2**). However, the observed difference in the average incidence of leptospirosis between the seasons of the year was not statistically significant.

**Table 2:** **Seasonal incidence of leptospirosis for the years 2010-2019, in Western Cape Province, South Africa.**

| **Year** | **Summer** | **Fall** | **Winter** | **Spring** |
| --- | --- | --- | --- | --- |
|  | IP/100,000<br>(95% CI) | IP/100,000<br>(95% CI) | IP/100,000<br>(95% CI) | IP/100,000<br>(95% CI) |
| 2010 | 0.10 (0.02,0.19) | 0.28 (0.14,0.41) | 0.12 (0.03,0.21) | 0.09(0.01,0.16) |
| 2011 | 0.12 (0.03,0.21) | 0.03 (-0.01,0.08) | 0.07 (0.001,0.13) | 0.10 (0.02,0.18) |
| 2012 | 0.07 (0.001,0.13) | 0.03 (-0.01,0.08) | 0.02 (-0.02,0.05) | 0.03 (-0.01,0.08) |
| 2013 | 0.05 (-0.01,0.10) | 0.05 (-0.01,0.10) | 0.07 (0.001,0.13) | 0.03 (-0.01,0.08) |
| 2014 | 0.02 (-0.01,0.05) | 0.10 (0.02,0.17) | 0.14 (0.05,0.24) | 0.19 (0.08,0.30) |
| 2015 | 0.20 (0.09,0.31) | 0.17 (0.07,0.27) | 0.16 (0.06,0.25) | 0.13 (0.04,0.21) |
| 2016 | 0.18 (0.08,0.29) | 0.21 (0.10,0.33) | 0.03 (-0.01,0.07) | 0.14 (0.05,0.23) |
| 2017 | 0.12 (0.03,0.18) | 0.09 (0.02,0.16) | 0.08 (0.01,0.14) | 0.06 (0.001,0.12) |
| 2018 | 0.12 (0.04,0.20) | 0.15 (0.06,0.24) | 0.09 (0.02,0.16) | 0.10 (0.03,0.18) |
| 2019 | 0.04(-0.01,0.09) | 0.07 (0.01,0.14) | 0.07 (0.01,0.14) | 0.10 (0.03,0.18) |
| **Average incidence** | **0.10 (0.03,0.18)** | **0.12 (0.04,0.20)** | **0.08 (0.02,0.15)** | **0.10 (0.02,0.17)** |

257 IP: incidence proportion; CI: confidence interval

### 3.3 Effect of sex, age, year, and season on leptospirosis incidence in WCP, 2010-2019

**Table 3** shows the results from the Multivariable Negative Binomial Regression (MNBR) model that was run to identify factors associated with the incidence of leptospirosis during the 10-year period. The incidence rate of leptospirosis among the male demographic was 2.2 (95% CI: 1.66,3.03) times higher than among females during the entire study period. The model results indicated that on average the incidence rate of leptospirosis among 18-44 and ≥ 45 age groups was 8.0 (95% CI: 4.65,15.15) and 7.4 (95% CI: 4.17,14.17) times higher than the incidence rate among ≤17 years age group. The incidence was substantially lower in 2012 (IRR 0.25; 95% CI 0.11-0.54), 2013 (IRR 0.34; 95% CI 0.16-0.68), 2017 (IRR 0.53; 0.28-0.98) and 2019 (IRR 0.47; 95% CI 0.24-0.87) compared to the incidence in 2010. From the model results, the incidence rate in fall, summer and spring seasons was 1.4 (95% CI: 0.95,2.15), 1.2 (95% CI: 0.78,1.81) and 1.2 (95% CI: 0.77,1.78) times higher than what was observed in winter season, however the observed difference was not statistically significant.

## 4. Discussion

This study provides a first description of the incidence of human leptospirosis in the Western Cape Province (WCP), South Africa across a 10-year period. Here we report an average incidence of 0.40 leptospirosis cases per 100,000 population, ranging from 0.15 in 2012 to 0.66 in 2015 cases per 100,000. There was no overall increase in the annual incidence of leptospirosis, however the incidence fluctuated widely across the 10-year period. The incidence was significantly higher in males compared to female. The 18-44-year-old and ≥ 45 years-old age groups had a higher incidence of the infection compared to the ≤17-year-old age group. The incidence did not vary based on season over the 10-year period.

**Table 3:** **MNBR analysis results between year, season, sex, and age group with incidence of leptospirosis**

| Variables | IRR | 95% CI | *P.value* |
| --- | --- | --- | --- |
| **Year of occurrence** |  |  |  |
| 2010 | Ref | — | — |
| 2011 | 0.543 | (0.281,1.026) | 0.061 |
| 2012 | 0.25 | (0.106,0.536) | **0.001** |
| 2013 | 0.337 | (0.157,0.684) | **0.003** |
| 2014 | 0.73 | (0.402,1.314) | 0.294 |
| 2015 | 1.033 | (0.597,1.796) | 0.907 |
| 2016 | 0.956 | (0.546,1.677) | 0.874 |
| 2017 | 0.53 | (0.282,0.98) | **0.046** |
| 2018 | 0.748 | (0.418,1.334) | 0.323 |
| 2019 | 0.466 | 0.244,0.872 | **0.019** |
| **Season of the year** |  |  |  |
| Winter | Ref | — | — |
| Spring | 1.166 | (0.765,1.782) | 0.475 |
| Summer | 1.184 | (0.778,1.808) | 0.431 |
| Fall/autumn | 1.422 | 0.947,2.147 | 0.091 |
| **Sex** |  |  |  |
| Female | Ref | — | — |
| Male | 2.239 | (1.664,3.034) | **<0.001** |
| **Age group** |  |  |  |
| ≤17 | Ref | — | — |
| 18-44 | 8.045 | (4.654,15.147) | **<0.001** |
| ≥45 | 7.391 | (4.167,14.169) | **<0.001** |

IRR: incidence rate ratio; CI: confidence interval; ref: reference category; MNBR: Multivariable Negative Binomial Regression.

The annual incidence observed during this 10-year period lies within the range of the incidence that has been reported globally to be occurring in temperate regions (0.1-1 cases per 100,000 population) [34]. The cold wet winters and warm dry summers might allow transmission and survival of pathogenic *Leptospira* species [21]. WCP is known for its harsh drought, and different types of floods such as river floods, flash floods and these occur in each year at varying intensities [35]. Annual fluctuation in such environmental conditions could partly explain the observed wide fluctuation in the annual incidence of leptospirosis because increase in incidence is normally reported after occurrence of extreme weather events such as heavy rains and high temperatures [29,34,36]. In addition, rodent infestation, increased prevalence of infected rodents and overcrowded settings were reported as risk factors for human leptospirosis outbreak in a one of the prisons in WCP in 2015 [22]. This outbreak could have contributed to the observed increase in the incidence in 2015. In a study conducted after this outbreak, Naidoo et al. further highlighted the importance of continued circulation of pathogenic *leptospira spp* among rodents in maintaining the status of leptospirosis transmission particularly in informal settlements [22].

Previous studies have indicated a significant relationship between leptospirosis incidence and seasonality [34,36,37]. In Western Cape, seasonality was not associated with the incidence of leptospirosis between 2010 and 2019, however the incidence was slightly higher in fall and summer as compared to winter and spring. This is consistent with some reports from other countries in temperate regions [34,38]. Studies on correlation between leptospirosis incidence and seasonality require consideration of other variables such as rodent seasonal population, temperature, rainfall, and other climate related parameters to be included in the analysis [37]. This could help in understanding the importance of the interaction of different environmental and climatic factors in determining the incidence of leptospirosis in resource limited settings.

The results showed that the overall incidence was significantly higher in males as compared to females (0.55 cases vs 0.25 cases per 100,000 population) during the study period. Similar results have been reported in both seroprevalence studies in Africa and in incidence studies conducted using surveillance data globally [26,27,34,38,39]. The higher incidence among males has been largely attributed to occupational or environmental exposure, whereby males engage in activities that may put them at higher risk of contracting the infection [36,40]. Epidemiological studies collecting data on occupation, underlying medical conditions, and place of settlement could aid in categorically establishing risk groups hence guiding policies on targeted prevention of leptospirosis.

The incidence of leptospirosis was highest among those within the 18-44 and ≥ 45 - year-old age groups compared to those below ≤17 years of age. These findings are in line with a systematic review by Costa et al. that reported the highest incidence to be occurring among adult males aged 20-49 years [5]. The observed results also correspond with other studies, where incidence has been shown to be higher among the active adult population [34,38]. People belonging to the 18-44 and ≥ 45 age groups would theoretically have an increased environmental exposure compared to those belonging to ≤17 age group, thus increasing their risk of being infected.

The data presented here are the first results to inform on the incidence trends and distribution of leptospirosis in WCP. The data covers a 10-year period, hence could help to guide designing of preventive strategies. However, there are some study limitations that need to be considered. Firstly, the study utilized data collected from passive surveillance and from a public health laboratory setting excluding data from private laboratories, hence there is a great potential for underestimation of the real occurrence of leptospirosis in WCP. Some bias may still be inherent in the analysis due to ambiguity in clinical presentation; the latter contributes to underestimation of actual incidence. Secondly, the analysis does not provide information on the geographical distribution of the cases within the province as such information was not included in provided data. Consideration of the geographical distribution of the incidence would have given insights into which specific districts are at risk, hence informing targeted interventions.

Findings from this analysis highlight the continued circulation of leptospirosis infection within WCP with age and sex being significant risk factors for infection. However, there is need of a good knowledge about the epidemiology of leptospirosis (such as the geographical and seasonal patterns, the specific risk populations, circulating *Leptospira* strains and the importance of reservoir animals) [13] within the province to improve prevention strategies, prediction, and detection of leptospirosis burden and outbreaks. For instance, future studies focusing on the impact of heavy rainfall, floods, seasonal fluctuation in weather-related factors, occupation, geographical location, and rodent dynamic parameters on leptospirosis incidence could help address the knowledge gaps on the actual burden of this emerging zoonosis.

In conclusion, the incidence of leptospirosis in WCP fluctuated within the range that has been reported to be occurring in temperate regions. The incidence was strongly related to sex and age; however, it did not differ across the seasons of the year. These results show that leptospirosis is an important zoonosis within the province and potentially disproportionately affecting males and the productive age demographic groups. The findings, therefore, can guide targeted intervention strategies within the province to decrease the burden of human leptospirosis.

## Data Availability

All data produced in the present work are contained in the manuscript

## 5. Acknowledgement

Special thanks to the National Health Laboratory Services (NHLS), South Africa for providing all the necessary data on leptospirosis tests conducted in Western Cape Province.

## 7. Supporting information

S1 Table 1: Summary of leptospirosis incidence distribution by age group from 2010 to 2019 in Western Cape Province, South Africa

S2 Table 1: Leptospirosis cases per healthcare facility between 2010-2019 in Western Cape Province, South Africa.

S3 Fig 1: University of Cape Town – Human Research Ethics Committee approval document.

