## Supplementary material for "Incidence and distribution of human leptospirosis in the Western Cape Province, South Africa, (2010-2019): A retrospective study": S1 Table 1

**S1 Table 1: Summary of leptospirosis incidence distribution by age group from 2010 to 2019 in Western Cape Province, South Africa**

| **Age group** | **≤17-year-old** | **18-44-year-old** | **≥ 45-year-old** |
| --- | --- | --- | --- |
|  | IP/100,000 (95% CI) | IP/100,000 (95% CI) | IP/100,000 (95% CI) |
| **Year of occurrence** |  |  |  |
| 2010 | 0.172 (-0.02,0.37) | 0.898 (0.54,1.26) | 0.521 (0.14,0.91) |
| 2011 | 0.113 (-0.04,0.27) | 0.366 (0.14,0.59) | 0.504 (0.13,0.88) |
| 2012 | 0 | 0.180 (0.02,0.34) | 0.279 (0.01,0.55) |
| 2013 | 0.218 (0.004,0.43) | 0.212 (0.04,0.38) | 0.135 (-0.05,0.32) |
| 2014 | 0.054 (-0.05,0.16) | 0.831 (0.50,1.16) | 0.196 (-0.03,0.42) |
| 2015 | 0.106 (-0.04,0.25) | 1.020 (0.66,1.39) | 0.632 (0.24,1.02) |
| 2016 | 0.052 (-0.05,0.15) | 0.570 (0.3,0.84) | 1.160 (0.64,1.69) |
| 2017 | 0 | 0.567 (0.30,0.84) | 0.298 (0.04,0.56) |
| 2018 | 0 | 0.525 (0.27,0.78) | 0.867 (0.43,1.31) |
| 2019 | 0 | 0.452 (0.22,0.69) | 0.337 (0.07,0.61) |
| **Mean Incidence** | **0.072 (-0.02,1.6)** | **0.562 (0.16,0.83)** | **0.493 (0.3,0.83)** |

IP; incidence proportion per 100,000 population: CI; confidence interval
