## Supplementary material for "Incidence and distribution of human leptospirosis in the Western Cape Province, South Africa, (2010-2019): A retrospective study": S2 Table 1

**S2 Table 1: Leptospirosis cases per healthcare facility between 2010-2019 in Western Cape Province, South Africa.**

| **District** | **Health Facility** | **n** | **%** |
| --- | --- | --- | --- |
| Cape Town Metro | Groote Schuur Hospital | 62 | 24.4 |
| Cape Town Metro | Mitchells Plain Hospital | 42 | 16.5 |
| Cape Town Metro | New Somerset Hospital | 34 | 13.4 |
| Cape Town Metro | Tygerberg Hospital | 27 | 10.6 |
| Cape Town Metro | Victoria Hospital | 26 | 10.2 |
| Cape Town Metro | Dietrich Voigt Mia TPY | 20 | 7.9 |
| Cape Town Metro | GF Jooste Trauma Hospital | 19 | 7.5 |
| Cape Town Metro | Khayelitsha Hospital | 10 | 3.9 |
| Cape Town Metro | 2 Military Hospital | 4 | 1.6 |
| Cape Town Metro | Helderberg Hospital | 3 | 1.2 |
| Cape Town Metro | Red Cross Children's Hospital | 2 | 0.8 |
| Cape Town Metro | Wynberg Clinic | 2 | 0.8 |
| Garden Route | George Hospital | 1 | 0.4 |
| Cape Town Metro | Karl Bremer Hospital | 1 | 0.4 |
| Cape Town Metro | Woodstock CDC | 1 | 0.4 |
|  |  | **254** | **100.0** |

n; frequency
